# Premature Cardiovascular Death and its Modifiable Risk Factors: Protocol of a Systematic Review and Meta-Analysis

**DOI:** 10.1101/2022.03.07.22272016

**Authors:** Wan Shakira Rodzlan Hasani, Nor Asiah Muhamad, Hasnah Maamor, Tengku Muhammad Hanis, Chen Xin Wee, Kamarul Imran Musa

## Abstract

**Introduction:** The burden of cardiovascular disease (CVDs), in the number of premature deaths, continues to increase globally and alarmingly, especially in almost all countries outside high-income countries. An accurate estimate of premature CVD death is of crucial importance for planning, implementing, and evaluating cardiovascular prevention and care interventions. However, existing literature reported the evidence on premature CVD death from selected regions, and there has been less evidence of systematic review with meta-analysis estimating the global premature death. This paper reports the protocol for a systematic review and meta-analysis to derive solid and updated estimates on global and setting-specific premature CVD death prevalence and its associated risk factors.

**Methods and analysis:** PUBMED, EMBASE, Web of Science, CINAHL, and Cochrane Central Register of Controlled Trials (CENTRAL) will be used as the literature database. Retrieved records will be independently screened by two authors and relevant data will be extracted from studies that report data on premature mortality (death before the age of 70 years) related to CVD. Study selection and reporting will follow the Preferred Reporting Items for Systematic Review and Meta-analyses (PRISMA) guideline. Pooled estimates of premature CVD mortality (based on standardised mortality ratio and years of life lost) and the effect size of modifiable risk factors will be computed applying random-effects meta-analysis. Heterogeneity among selected studies will be assessed using the I^2^ statistic and explored through meta-regression and subgroup analyses. Depending on data availability, we propose to conduct subgroup analyses by geographical area, CVD events, and socio-demographic variables of interest study. The risk of bias for the studies included in the systematic review or meta-analysis will be assessed by the Newcastle–Ottawa Quality Assessment Scale.

**Ethics and dissemination:** Ethics approval is not required as the data used in this systematic review will be extracted from published studies. The systematic review will focus on the premature CVD mortality rate and its associated factors. The findings of the final report will be disseminated to the scientific community through publication in a peer-reviewed journal and presentation at conferences.

**PROSPERO registration number:** **CRD42021288415**

## INTRODUCTION

Globally, cardiovascular diseases (CVDs) contributed to the most non-communicable disease (NCDs) death (17. 9 million people) followed by cancers (9.3 million), respiratory diseases (4.1 million), and diabetes (1.5 million). These four groups of diseases represent more than 80% of all premature NCD deaths [1]. Premature death is a death that occurs before the average age of death in a certain population [2]. It was measured in terms of potential years of life lost (PYLL) before the age of 70 years [3]. PYLL is an estimate of the average years a person would have lived if she or he has not died prematurely [4]. It was a standardised parameter commonly used to measure the burden of disease due to premature mortality or death. According to World Health Organization (WHO), the burden of premature death was notably high in the low- or middle-income countries (LMICs) where over three-quarters of premature deaths occur in LMICs [1].

The most comprehensive global estimate of mortality research, theGlobal Burden of Disease (GBD) study reported ischemic heart disease (IHD) and stroke remain the leading causes of premature death worldwide in 2010 [5]. According to GBD data, the number of CVD deaths steadily increased from 12.1 million in 1990 to 18.6 million in 2019 where the global trends for disability-adjusted life years (DALYs) and years of life lost due to IHD and stroke also increased significantly [6].

Previous literature reported cardio-metabolic risk, behavioural risk, environmental, and social risk factors contributed to CVD death [7-10]. A multi-center study involving 21 countries from the middle, low, and high-income countries by Yusuf et al., demonstrated the CVD deaths in the overall cohort were attributed to modifiable risk factors [11]. They found diabetes was the strongest risk factor for death among metabolic factors, followed by hypertension and abdominal obesity. Whereas for behavioural risk, tobacco use demonstrated the strongest association with death, followed by high alcohol consumption, low physical activity, and poor diet. The global estimates of the associations between risk factors and adult deaths from GBD data also reported CVD burden attributable to modifiable risk factors continues to increase globally [6].

### Why is it important to do this review?

In line with the Sustainable Development Goal (SDG) 3 targeting a 30% reduction in premature mortality due to NCDby 2030, there is an urgent need to focus on revising the existing policies for the prevention and control of chronic diseases [12]. Information on premature CVD mortality and its associated modifiable risk factors may assist the development of global and context-specific strategies for reducing the incidence of premature CVD mortality. However, the most widespread global estimates of risk factors for CVD death by the GBD study [13, 14] are derived through combining data with relatively little data from LMICs. To date, there is an absence of a comprehensive systematic review with a meta-analysis study that has analytically explored the association of common modifiable risk factors and premature CVD death globally. Thus, we aim to conduct a systematic review of the studies on premature CVD mortality and perform a meta-analysis to estimate the global and setting-specific (e.g. difference geographical/continent and subgroup population) of premature mortality related to any CVD event.

### Objective

This protocol is designed to establish and synthesise information on premature CVD mortality among adults below 70 years old. We will explicitly describe the methodology for conducting a systematic review and meta-analysis for this review. We will systematically review all published evidence to determine the pooled estimates of premature CVD mortality using the standardised mortality ratio (SMR) and years of life lost (YLL). Subsequently, subgroup analyses will be conducted to determine the premature CVD mortality by geographical areas (six continents including Asia, Africa, Europe, North America, South America, and Oceania), two main CVD types (ischemic heart disease and cerebrovascular disease), and gender. We will also determine the commonest modifiable risk factors (or the highest risk ratio) associated with premature CVD mortality.

### Review questions

For patients below 70 years who are confirmed died due to CVD event, we will determine:

1. what is the pooled estimate premature CVD mortality, in overall and according to the geographical area (six continents), CVD events, and gender?
2. what are the important modifiable risk factors associated with premature CVD mortality that has been identified in previous studies?

## METHOD

This systematic review will be reported and conducted following the Preferred Reporting Items for Systematic Reviews and Meta-analysis (PRISMA) protocol [15]. Simultaneously, we will integrate this report with theMeta-analysis of Observational Studies in Epidemiology (MOOSE) guideline [16] since the outcome of this review will be meta-analysis from selected observational studies. This protocol was registered with the International Prospective Register of Systematic Reviews (PROSPERO), [Registration number: CRD42021288415].

### Patient and public involvement

Patients and the public will not be involved in this study as this is a systematic review protocol.

### Eligibility criteria

#### Types of studies

This systematic review will review published articles restricted to original research papers that use an observational study design (including cohort, case-control, and cross-sectional studies) that report premature mortality due to CVDs or any intervention studies which focus on prevention or treatment for premature CVD mortality. We will exclude any review, case study, commentaries, or qualitative studies.

#### Population

This review will include all premature mortality among adults below the age of 70 years due to CVD (including coronary heart disease, cerebrovascular disorder, myocardial ischemia or stroke). We choose 70 years as the upper limitfor premature death as recommended by WHO due to the increasing level of co-morbidity and uncertainties of the estimation of cause-specific death rates at older age [17]. The outcomes of CVD events in this review refer to diseases reported based on the International Classification of Diseases (ICD-10) codes – (i) I20-I25 for ischemic heart diseases and I60-I69 for cerebrovascular diseases.

#### Types of exposure

We will include studies reporting premature CVD mortality outcomes or investigating modifiable risk factors associated with premature CVD mortality including cardio-metabolic, and behavioural such as diabetes, hypertension, hypercholesterolemia, obesity, tobacco smoking, alcohol use, unhealthy diet, physical inactivity or low socioeconomic (low income, low education level and employment status) either as a single factor or multiple risk factors.

#### Types of outcome measures

The outcome for this review is premature mortality due to CVD event. Confirmed premature CVD mortality is defined by a description of clinical signs or history where deaths occur before life expectancy (<70 years) is reached. We plan to determine the pooled estimate of premature CVD mortality using a standardised mortality ratio (SMR). The SMR gives the ratio of death that is due to CVD compared to the general population. For each cause of death, SMRs and their 95% confidence intervals will be extracted from each publication. If not reported, SMR will be calculated using this formula:

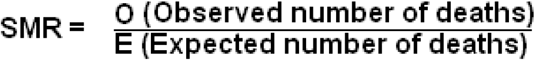

Another key parameter in measuring premature mortality is years of life lost (YLL). Thus, the pooled estimates of YLL due to CVD events will be estimated in this analysis. The method of calculating YLL will be collected from related articles. We will include the original method of Murray and Lopez for YLL calculation, where the number of deaths was multiplied by standard life expectancy for each age group and summed for all age groups [18]. The analysis of pooled premature CVD mortality using SMR and YLL will be stratified according to the two commonest CVD events (ischemic heart disease and cerebrovascular disease), geographical area (six continents), and gender whenever data is available. We will also tabulate the effect size (measured in risk ratio) of the modifiable risk factors associated with premature CVD mortality that fulfil the inclusion criteria for this review. The important risk factor with the highest risk ratio associated with premature CVD mortality will be determined.

### Information sources

#### Electronic search

We will systematically conduct a comprehensive literature search using various literature databases, including PubMed, EMBASE, Web of Science (WoS), Cumulative Index to Nursing and Allied Health Literature (CINAHL), and Cochrane *Central* Register of Controlled Trials (*CENTRAL*) to identify eligible studies. In addition, secondary searches in other sources, such as Google Scholar and grey literature (such as unpublished studies) will also be carried out. The reference section of the included studies also will be hand-searched for additional relevant studies.Studies will be restricted to the English language and there is no restriction on the publication date.The search will be performed from selected electronic databases up to April 2022.

#### Search strategy

Table 1 (see Appendix 1) shows the proposed search term for search strategy. The first theme is “cardiovascular diseases”. The second theme is “modifiable risk” or “behavioural risk” including hypertension, hyperglycemia, hyperlipidaemia, obesity or overweight, tobacco use, physical inactivity, unhealthy diet, alcohol use, and low socioeconomic status. The third theme is “premature mortality” including “years of life lost” and “standardise mortality ratio” and the last theme is population (young/adults/middle age). The exploded versions of Medical Subject Headings (MeSH) of each theme will be included. All the four search themes will be combined using the Boolean operator ‘AND’.

### Study selection

Two review authors (WS and NAM) will independently screen all the titles and abstracts to examine the potential studies for inclusion. We will identify the studies and will code them as ‘retrieve’ (eligible or potentially eligible/unclear) or ‘do not retrieve’. We will retrieve the full-text study reports/publications, and the review authors (WS and NAM) will independently screen the full text to identify studies for inclusion, as well as identifying and recording reasons for exclusion of the ineligible studies. We will resolve any disagreement through discussion or, whenever required, we will consult a third review author to make the final judgment (KIM).

We will identify and exclude duplicates and collate multiple reports of the same study so that each study rather than each report is the unit of interest in the review. We will record the selection process in sufficient detail to complete a PRISMA flow diagram (Figure 1) and construct a table describing the characteristics of the excluded studies [15, 19]. The bibliographic software EndNote (https://www.myendnoteweb.com/) [20] will be used to store, organize, and manage all the references and ensure a systematic and comprehensive.

**Figure 1:**
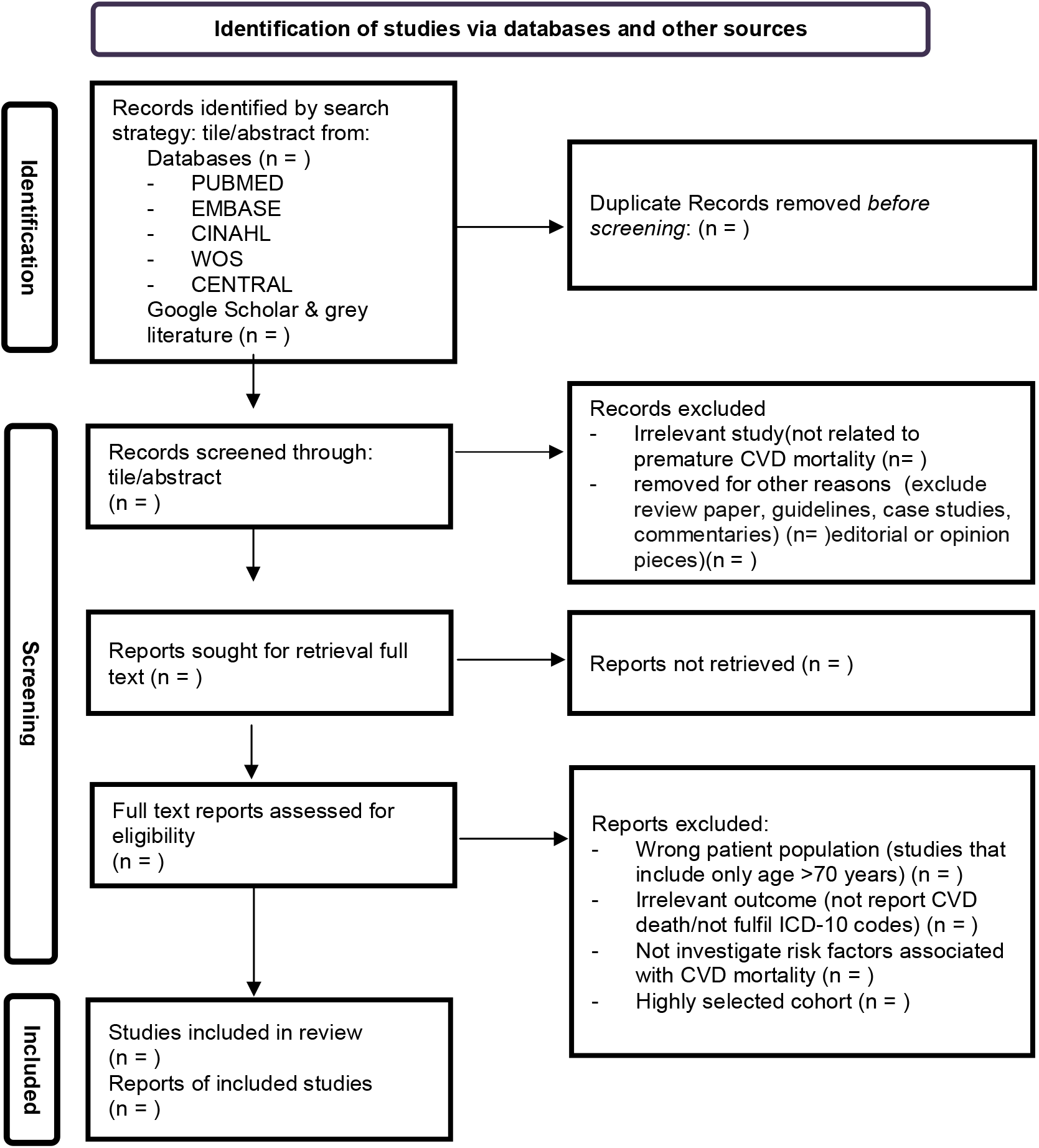
Flow diagram of studies that were considered for inclusion.

#### Data extraction and management

We will use a standardised data extraction form created by the Microsoft Excel Spreadsheet Software for study characteristics and outcome data.One review author (WS) will extract the following study characteristics from the included studies; -

- Title, authors, and publication year
- Study objectives
- Methods: study design, total duration of the study, study location, study setting, study period, and method of analysis.
- Participants: N, mean age or age range, gender, the severity of the condition, diagnostic criteria if applicable, inclusion criteria, and exclusion criteria.
- Exposures or risks: description of exposure risks such as types of CVD: either ischemic heart disease or cerebral vascular accident (ischemic or haemorrhagic), duration, intensity, medication if applicable, and control condition.
- Outcomes: description of primary and secondary outcomes specified and collected, and time points reported.

Two review authors (WS, NAM) will independently extract outcome data from the included study. We will note in the ‘Characteristics of included studies’ table if outcome data is not reported in a usable way. We will resolve any disagreements by consensus or by involving a third review author (KIM). We will double-check that data is entered correctly by comparing the data present in the systematic review with the study reports.

#### Quality assessment

Two review authors (WS, NAM) will independently assess the study quality according to the criteria incorporated from Newcastle-Ottawa Scale (NOS) which is the tool for quality assessment for non-randomized studies and meta-analysis[19]. We will include studies with a potentially high risk of bias. We will assign each potential source of bias as high, low, or unclear and provide a quote from the study report together with justifications for our assignment in the quality assessment in included studies table.NOS applied a ‘star system’ where the study is assessed based on three broad perspectives: 1) the selection of the study groups; 2) the comparability of the groups, and 3) the ascertainment of exposure/outcome. The maximum score is 9 points, where the studies can be classified as good, fair or poor quality according to following standard thresholds; -

1. Good quality: 3 or 4 stars in selection domain AND 1 or 2 stars in comparability domain AND 2 or 3 stars in outcome/exposure domain
2. Fair quality: 2 stars in selection domain AND 1 or 2 stars in comparability domain AND 2 or 3 stars in outcome/exposure domain
3. Poor quality: 0 or 1 star in selection domain OR 0 stars in comparability domain OR 0 or 1 stars in outcome/exposure domain.

### Summary of findings table

A table will be created to summarise the characteristics of the included studies. The primary outcome of this review will be the arrays of premature mortality due to CVD events associated with modifiable risk factors. Hence, we will create a ‘summary of findings’ table by tabulating the SMR and YLL following with modifiable risk factors: (1) diabetes, (2) hypertension, (3) hypercholesterolemia, (4) obesity, (5) smoking, (6) alcohol use, (7) unhealthy diet, (8) physical inactivity and (9) socio-economic factors.

### Data analysis and Statistical analysis

Statistical analysis will be conducted using R software. The main R packages, “meta”[21] and “metafor” [22] will be used for meta-analysis. When data are available, we will tabulate the count, proportion, incidences, or mortality ratio of premature CVD mortality. The random effect model will be used in meta-analysis to calculate the pooled estimates of premature mortality based on the SMR and YLL.The analysis will be stratified according to two commonest CVD events, geographical area (six continents), and gender whenever possible. The assessment of heterogeneity will be done using the I^2^ statistic and visual inspection of Forest plots. We will use risk ratio (RR) for dichotomous exposure measures and will present with the corresponding 95% CIs. If the results cannot be entered as detailed here, we will describe them in the ‘Characteristics of included studies’ table or enter the data into additional tables.

### Assessment of heterogeneity

We will visually inspect the data to check for the presence and identify the sources of heterogeneity among the studies. We will decide whether it is sensible to combine the studies in a single meta-analysis based on the similarity of population, exposure, and outcome. We will consider populations as similar when they are exposed to similar potential aetiological factor(s) for premature mortality, with outcomes measured in similar ways, as stated above in ‘Types of outcome measures’. We plan to estimate the exposure effects of individual studies and examine heterogeneity between studies by inspecting the forest plots and quantifying the impact of heterogeneity using the I^2^ statistic. For statistical heterogeneity, we will explore the possible causes (for example, differences in study quality, participants, exposures, or outcome assessments). For a moderate or high degree of heterogeneity, we will evaluate the studies in terms of their methodological characteristics to determine whether the degree of heterogeneity can be explained by differences in those characteristics and whether a meta-analysis is appropriate.

#### Assessment of reporting biases

We will create and examine a funnel plot to explore possible small study biases if we can pool more than ten studies in a single meta-analysis. The number of studies that are missing from the funnel plot will be estimated. The effect size after the imputation of these missing studies will be estimated by the trim-and-fill method. The trim and fill method is a simple estimation approach proposed by Duval and Tweedie[23] where they trim off the asymmetric outlying part of the funnel, then use the symmetric remainder to estimate the true center of the funnel and then replace the trimmed studies and their counterparts around the centre. Other methods to assess the publication bias including the usage of Bregg’s rank correlation [24] and Egger’s weighted regression methods test [25] will also be performed.

## DISCUSSION

The increasing cases of premature death due to CVD warrant the researchers to obtain the updated analysis on the global premature mortality rate (or prevalence of premature death) of CVD. Limited search by the authors revealed no recent (for the past five-years) meta-analysis thus far focusing on premature death due to CVD events.We will base our conclusions only on findings from the quantitative synthesis of included studies for this review. We strongly believe that our conclusion will be of crucial importance for both research and future policy-making, especially assist in formulating a more effective and targeted non-communicable disease prevention strategies. Our finding also might inform the development of mathematical models or cost-effectiveness analysis to get projections of future premature death burden due to CVD.The data might help to identify settings or subgroups of the population where the risk for CVD death is of higher concern and needed special prevention priorities (e.g., in a specific continent/gender-based interventions). Future qualitative studies could be conducted to explore the modifiable determinants of CVD in certain high-risk subgroup/settings.

Strengths and limitations will be highlighted in the process of identifying evidence. This systematic review will yield solid and updated estimates on the global prevalence of premature CVD mortality. Unlike previous global reports, data included will not be limited to studies conducted in high-income countries, and records from LMICs will be included. The meticulous design, use of standardised study rating instruments, and compliance with all relevant guidelines for systematic reviews and meta-analyses are the anticipated strengths. Limitations will mainly originate from different designs and characteristics of all the included studies; this may lead to high heterogeneity which will, in turn, lower the quality of the evidence of this meta-analysis and systematic review; however, this may be overcame by including subgroup analyses and meta-regression in the meta-analysis.

## Data Availability

All data produced in the present study are available upon reasonable request to the authors

## ETHICS AND DISSEMINATION

We registered this systematic review with the National Medical Research Register, Ministry of Health Malaysia. There will be no concerns about privacy. The systematic review will focus on premature CVD mortality rate and its associated factors, the results of which will be disseminated by publication in a peer-reviewed journal after completion.

## Acknowledgments

We would like to thank the director general of Health Malaysia for allowing us to publish this report. We would also like to express our gratitude to the Universiti Sains Malaysia and the National Institutes of Health Malaysia for their support and technical guidance in developing this protocol.

## Patient consent for publication

Not required.

## Appendix 1

**Table 1:**
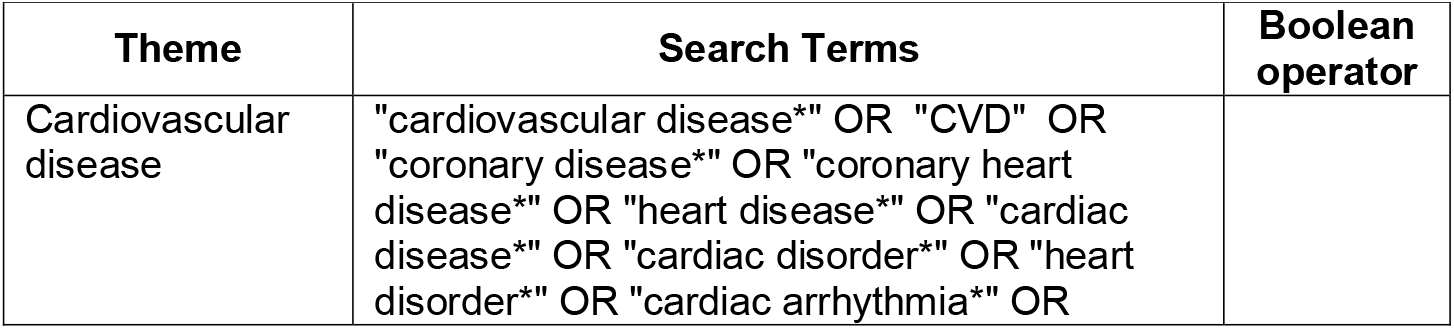
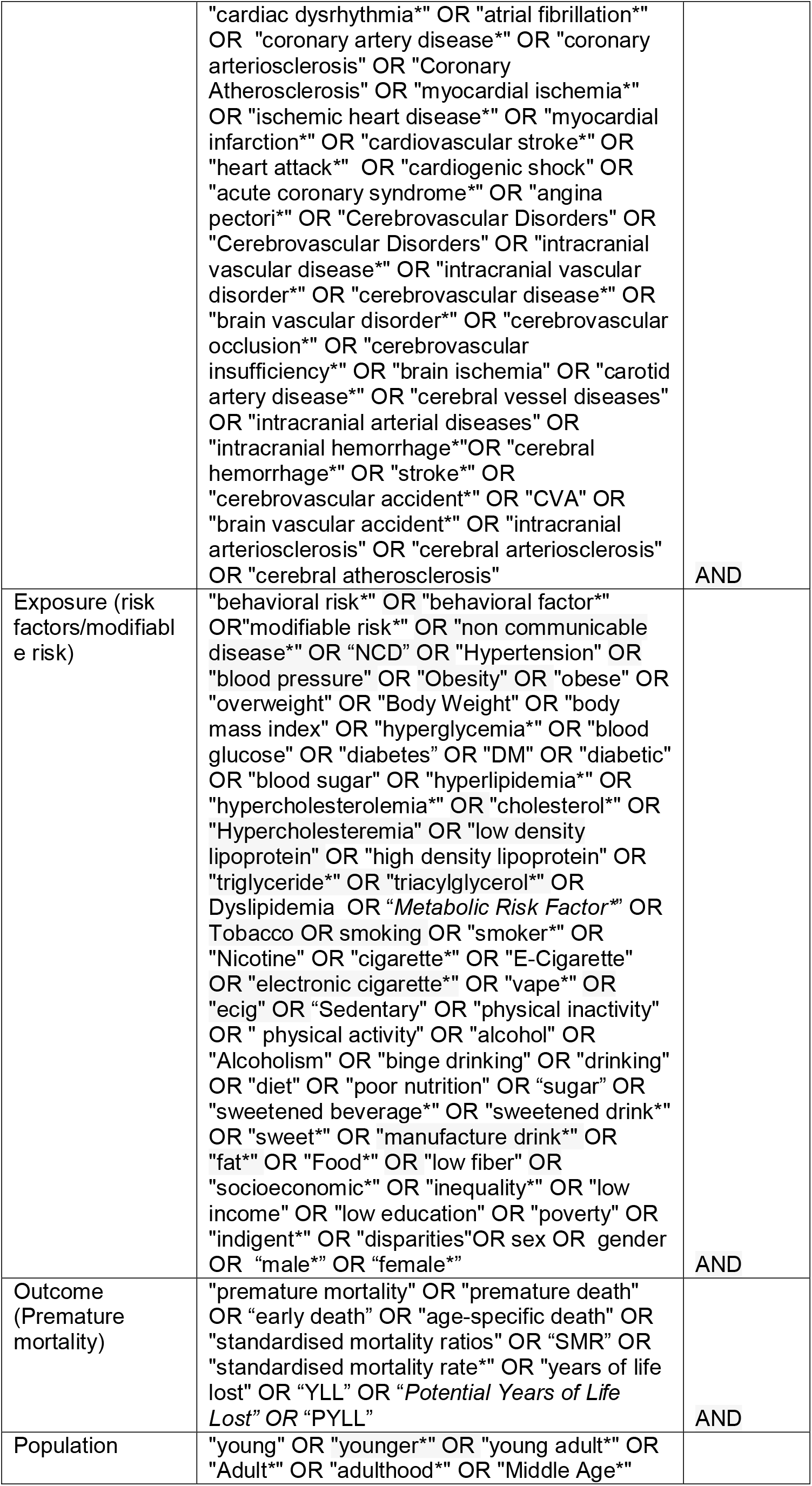
Propose search term.

